# Leveraging the U.S. blood supply to detect emerging viral threats

**DOI:** 10.64898/2026.02.06.26345722

**Authors:** Lennart J. Justen, Harmon Bhasin, Charles Whittaker, Daniel Cunningham-Bryant, Marion C. Lanteri, Michael P. Busch, Kevin M. Esvelt, Pardis C. Sabeti

## Abstract

Emerging infectious diseases often circulate undetected until they cause clinical illness, leaving surveillance systems reactive rather than anticipatory. The U.S. blood supply, with millions of whole-blood and plasma donations annually, offers an underutilized national resource for proactive pathogen surveillance. We propose integrating metagenomic sequencing (MGS) into existing blood collection and testing workflows to detect known and novel viruses in residual samples within days of collection. Unlike current blood screening, which targets a limited set of predefined pathogens, MGS captures a broad range of viral threats, including blood- and vector-borne agents that produce asymptomatic viremia and may be missed by syndromic surveillance. Modeling suggests that an annual investment of $5.5 million could detect a novel HIV-like pathogen before it infects 0.01% of the U.S. population. Building on established donor monitoring, testing, and privacy infrastructure, blood supply sequencing represents a deployment-ready opportunity to strengthen national and global biosecurity.

## 1. Introduction

Early detection of emerging infectious diseases is essential to protect public health and national biosecurity, yet most detection systems remain reactive rather than proactive. They are designed to detect familiar pathogens or clinical illness, not the early circulation of novel threats, particularly those that spread silently through long incubation periods and asymptomatic infection. Such delays have defined some of the most consequential epidemics, including HIV, hepatitis B (HBV), and hepatitis C (HCV), each of which circulated for years before it was recognized and controlled. Preventing a similar failure today requires scalable systems that can detect new pathogens before they trigger outbreaks.

The U.S. blood supply is well positioned to support this kind of early warning. Approximately 12 million whole-blood donations and 40 million plasma donations are collected each year,^1,2^ with the majority of transfusion-transmitted infection (TTI) testing consolidated within a small number of high-throughput laboratory networks. Aliquots from individual whole blood donations are routinely pooled for nucleic-acid testing (NAT), with residual material often available within 48 hours of collection. This provides standardized, rapid access to quality-controlled samples spanning a demographically and geographically distributed population. Aggregating pathogen information across millions of donors combines some of the strengths of environmental surveillance approaches such as air^3^ and wastewater^4^ with those of individual sampling, including greater sensitivity and direct evidence of human-to-human transmission.^5^

Current blood-supply testing does not, however, support detection of emerging pathogens. TTI screening uses targeted nucleic-acid and antibody assays to confirm the presence or absence of roughly a dozen pre-specified agents.^6,7^ TTI testing is central to transfusion safety and has substantially reduced transmission risk, but it is not designed to detect pathogens outside its target panel. This structural limitation represents an ongoing vulnerability in transfusion safety, and also a missed opportunity to leverage the blood supply for pathogen early warning. HIV, HBV,^8^ HCV,^9^ and human T-lymphotropic virus (HTLV)^10^ circulated through the blood supply and the general population for years before each was identified. HIV infected an estimated 12,000 transfusion recipients in the U.S. between 1978 and 1984,^11^ and nearly 25% of transfusion recipients were infected with HCV in the decade before screening began in the early 1990s.^9^

Metagenomic sequencing (MGS) of blood supply samples would strengthen pathogen early warning and transfusion safety by sequencing all nucleic acids in a sample without prior targeting, enabling detection of both known and novel pathogens in a single assay^12^. MGS has been used repeatedly to discover novel viruses^13^ and identify unexpected or fastidious pathogens, including from blood samples.^14–18^ Prior work has proposed MGS as an enhancement to transfusion safety and demonstrated one-off efforts to characterize the virome of blood donors.^19–22^ This paper develops the blood supply’s underexplored potential as an integrated pathogen early warning platform.

Routine MGS-based monitoring of the blood supply is practical today in a way it was not when first proposed two decades ago.^23^ Sequencing costs have fallen by several orders of magnitude, bringing nationwide deployment within economic reach. Decades of National Heart, Lung, and Blood Institute (NHLBI)-funded research through the Recipient Epidemiology and Donor Evaluation Study (REDS, 1989–present)^24,25^ and the FDA’s Transfusion-Transmitted Infections Monitoring System (TTIMS, 2015–present)^26^ have established the partnerships, sample processing pipelines, and data-sharing frameworks that a national surveillance program would need. During the COVID-19 pandemic, the REDS-IV program rapidly launched the RESPONSE study to screen blood mini-pools for viral RNA^27^ and later partnered with the CDC to analyze more than 1.4 million donor specimens across all 50 states for SARS-CoV-2 antibodies.^28,29^ As national biosecurity and public health strategies increasingly prioritize early detection of novel pathogens, the blood supply stands out as a powerful, implementation-ready sample stream.^30,31^

We estimate that an annual investment of approximately $5.5 million (range $3.5–$26.2 million) could support a national MGS-based blood monitoring program capable of detecting a novel HIV-like pathogen before it infects 0.01% of the U.S. population. This paper lays out what such a system would look like in practice, focusing on the U.S., although the approach is broadly applicable to countries with comparable blood collection infrastructure. We describe the MGS approach, map the operational pathways for sampling within the U.S. blood and plasma supply, estimate implementation costs, assess which pathogen types are most detectable in asymptomatic donors, and review the privacy and regulatory frameworks that apply to secondary sample use.

## 2. Monitoring approaches within the U.S. blood supply

The blood supply comprises two main components, both of which can be leveraged for MGS-based pathogen surveillance with rapid turnaround times and minimal disruption to existing operations. The transfusion medicine sector collects whole blood from unpaid donors, and the plasma industry collects plasma from paid participants for the manufacturing of plasma-derived medicinal products (PDMPs) (see McCullough^32^ for a detailed overview). Both sectors are regulated by the FDA’s Center for Biologics Evaluation and Research (CBER), which enforces strict donor eligibility and health requirements^33^ to ensure donations primarily originate from asymptomatic individuals,^32^ and mandates testing for known TTIs.^34^ Much of this testing is already centralized through nonprofit laboratory organizations, most notably Creative Testing Solutions (CTS), which tests approximately 60% of all U.S. blood and plasma donations annually.^35^

An MGS-based system would analyze residual samples generated during routine collection and testing. Key sample streams include leftover aliquots from individual or pooled TTI testing,^28,36^ donation units that test positive for TTIs,^34^ and large plasma manufacturing pools.^37^ These samples could be sent to a designated laboratory for sample processing and MGS, with bioinformatic analysis^13^ guiding follow-up investigations as needed, including deployment of targeted assays to validate potentially concerning detections. MGS complements rather than replaces these targeted assays. Targeted assays are generally more sensitive for known agents and are clinically validated under FDA regulations; MGS is not yet a regulated diagnostic technology and presents distinct challenges around contamination control and bioinformatic implementation, including reference database curation.^38^ We focus here on a near-term implementation in which MGS is applied to pooled, de-identified residual samples for pathogen monitoring and early warning, with confirmatory transfusion safety testing performed through existing TTI traceback infrastructure when warranted.

### 2.1 Whole blood collection for transfusion medicine

In the U.S., the transfusion medicine sector primarily collects whole blood—unseparated blood containing plasma, red blood cells, white blood cells, and platelets—from voluntary, unpaid donors for direct therapeutic use or component separation.^32^ The scale is substantial: in 2021, 6.6 million U.S. donors provided 11.5 million donations,^1^ offering broad population representation. Collection occurs through a distributed network of community-based organizations and hospital collection centers.^39^ Most community-based collection is managed by the American Red Cross (ARC, ∼40% of supply) and members of America’s Blood Centers (ABC, ∼60%),^40^ including large organizations such as Vitalant.

Standard TTI testing procedures generate samples well suited for MGS. During donation, multiple aliquots, typically two to three tubes totaling 20-30 mL, are collected concurrently with or immediately following the main donation unit for TTI testing.^32,34^ These are processed into plasma or serum and preserved with anticoagulants such as EDTA to prevent clotting and maintain nucleic acid integrity. To improve testing efficiency, blood centers often pool samples from 4-24 donations into “mini-pools” (MP)^27^ for initial nucleic acid testing (MP-NAT) to detect viruses such as HIV, HCV, and HBV. If a pool tests positive, individual samples are subsequently tested to identify the infected donor(s). This workflow yields residual individual donor aliquots, mini-pools, and TTI-positive units available for MGS pathogen surveillance (Figure 1), with the potential to further pool any samples prior to sequencing. Residual mini-pools are the most promising primary stream, as they already aggregate contributions across multiple donors, enhancing donor privacy and cost-efficiency, and are continuously generated by existing TTI testing at scale.

**Figure 1.**
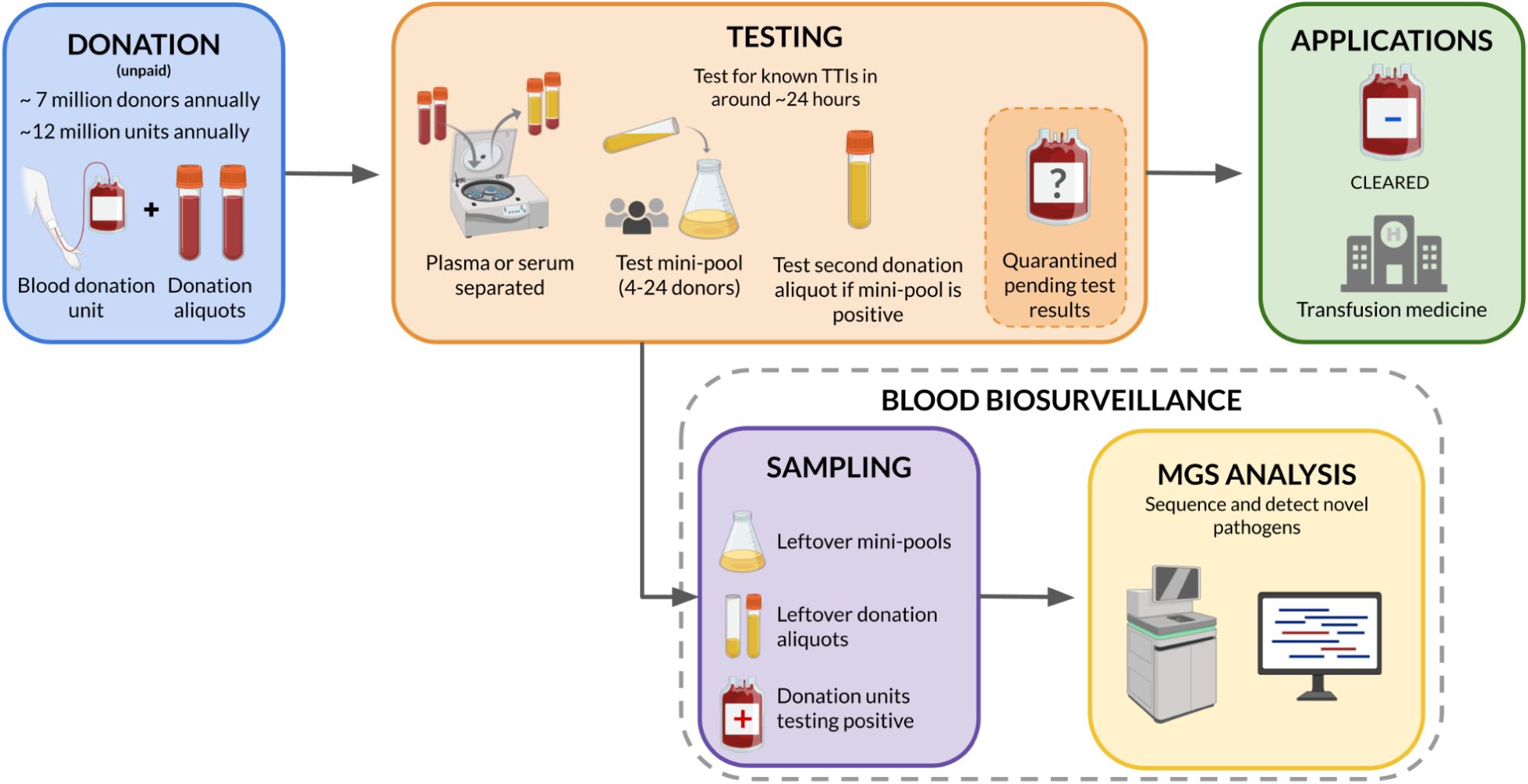
Sampling strategies in the transfusion medicine sector for metagenomic sequencing (MGS)-based biosurveillance. The standard whole blood donation and testing process generates multiple sample streams, including leftover donation aliquots, mini-pools, and transfusion-transmitted infection (TTI)-positive units, that can be repurposed for MGS and pathogen-agnostic detection with minimal operational disruption.

Rapid access to these residual samples is feasible. Operational constraints, including the short five-day shelf-life of platelets,^32^ necessitate rapid TTI testing, with results often reported within 24 hours of donation.

Previous efforts leveraging the blood supply for pathogen monitoring have established a strong foundation for accessing and analyzing these samples.^27,29,41,42^ During the COVID-19 RESPONSE study, approximately 3,000 residual mini-pools from blood donor testing were sent to research laboratories each month for SARS-CoV-2 NAT, ultimately encompassing more than 250,000 donations over the study period.^27^ Other initiatives included hepatitis E virus (HEV) RNA and seroprevalence testing using residual samples from over 18,000 routine blood donations obtained from the ARC,^41^ and Vitalant’s testing of individual donor aliquots from more than 500,000 unique donors across 800,000 donations for SARS-CoV-2 antibodies.^42^ These and other examples illustrate that large-scale access to de-identified residual samples through partnerships between blood collection organizations, public health agencies, and research institutions is feasible and well-established,^26,43^ and could readily support a nationwide MGS-based surveillance system.

### 2.2 Source plasma collection for plasma-derived medicinal products (PDMPs)

The plasma industry collects “source plasma”—plasma obtained via plasmapheresis from compensated donors^44^— for manufacturing PDMPs.^45^ This for-profit sector is vast, with 3.1 million individuals contributing plasma in the U.S. in 2019^46^ and 43 million donation events in 2023.^2^ Compared to whole blood donors, who are often majority Caucasian, source plasma donors tend to be younger, more frequently male, and include a higher representation from communities of color.^47^ Source plasma can also be donated more frequently than whole blood; up to twice weekly compared to every eight weeks.^48^ The U.S. supplies roughly 70% of the world’s plasma for manufacturing.^49^ The market is highly concentrated, with a few vertically integrated multinational companies (CSL Plasma, Grifols, Biolife Plasma by Takeda, and Octapharma AG) operating over 85% of U.S. plasma centers.^46^

As in transfusion medicine, the plasma industry’s TTI testing workflows present several opportunities for MGS. Key sample streams generated during routine operations include leftover donation aliquots, mini-pools, and TTI-positive units. Individual samples for TTI testing are drawn as separate aliquots during donation, recovered from collection tubing, or dispensed directly from the donation unit after collection. Donation aliquots are pooled for MP-NAT, utilizing mini-pools with pool sizes ranging from 50 to 1,000 samples.^50^ As in the transfusion medicine sector, residual minipools are the most promising primary stream, aggregating substantially more donors per pool, further improving cost-efficiency and donor privacy.

Another potential sampling stream arises from plasma fractionation–the separation of plasma into different therapeutic components–, where plasma from thousands of donors (typically 1,000 to 10,000, but sometimes up to 60,000) is combined into large manufacturing pools.^37,51^ These pools have been used for MGS in prior studies,^37^ but their value for early pathogen detection is limited by a mandatory 45- to 60-day inventory hold period on all plasma units^44^ instituted to allow time for disqualifying donor information to surface. Their delayed availability makes them less suited to early warning, but well suited to characterization of the broader donor virome and to retrospective investigation of suspected outbreaks.

Implementing MGS surveillance within the plasma industry requires collaboration with private companies. Plasma companies like Grifols have performed viral serosurveillance using residual source plasma testing aliquots pooled across more than 2,500 U.S. donors.^52^ Researchers have also conducted MGS on samples taken from four U.S. plasma manufacturing pools with around 2,500 donations per pool, on average,^37^ demonstrating the feasibility of accessing diverse sample types from this industry. These precedents indicate that integrating MGS into plasma collection and testing workflows is operationally tractable and would extend blood-based pathogen surveillance to a much larger and more frequently sampled population.

## 3. Modeling the costs of blood supply monitoring

To estimate the cost of an MGS-based blood monitoring system, we developed a modeling framework to calculate the annual investment required to detect an emerging pathogen before it exceeds a specified cumulative incidence target (e.g., 0.01% of the population infected). The model, adapted from prior work,^53^ integrates outbreak dynamics, pathogen characteristics, and sequencing parameters to determine the weekly sequencing depth necessary for detection with 95% probability. This depth requirement is then translated into an annual cost by incorporating sample acquisition, lab processing, data management, and personnel expenses. Full details of the model structure and assumptions are provided in the Appendix.

We applied this framework to a case study of a hypothetical novel, HIV-like pathogen. This simulated virus was modeled with a slow exponential growth rate (doubling time of around 43 weeks) and a 12-week period of continuous viremia following infection. The mean relative abundance of pathogen reads in infected individuals was set at μ=1.26×10^-5^, based on clinical MGS data from early-stage acute HIV infections.^54^ The detection threshold was defined as the accumulation of 100 reads corresponding to the target pathogen. In our central scenario, we assume that infected individuals are equally likely to donate as uninfected individuals, but we explore the sensitivity of our results to reducing the chance of an infected individual donating relative to an uninfected individual.

Our analysis highlights a strong dependence of required sequencing depth on detection timing and a clear trade-off between earlier detection and cost. As shown in Figure 3b, annual expenses to detect a novel HIV-like pathogen range from roughly $1.4 million at a cumulative incidence of 0.1% to just over $61 million at 0.001%. At lower cumulative incidence detection targets, the pathogen’s rarity in the population necessitates larger weekly cohorts to ensure that infected individuals are captured for sequencing, driving up both sampling and sequencing costs. Figure 3c highlights that reducing the chance of an infected individual donating relative to an uninfected individual increases the required sequencing depth, though the effect is modest for relative risks above 1/5.

**Figure 2.**
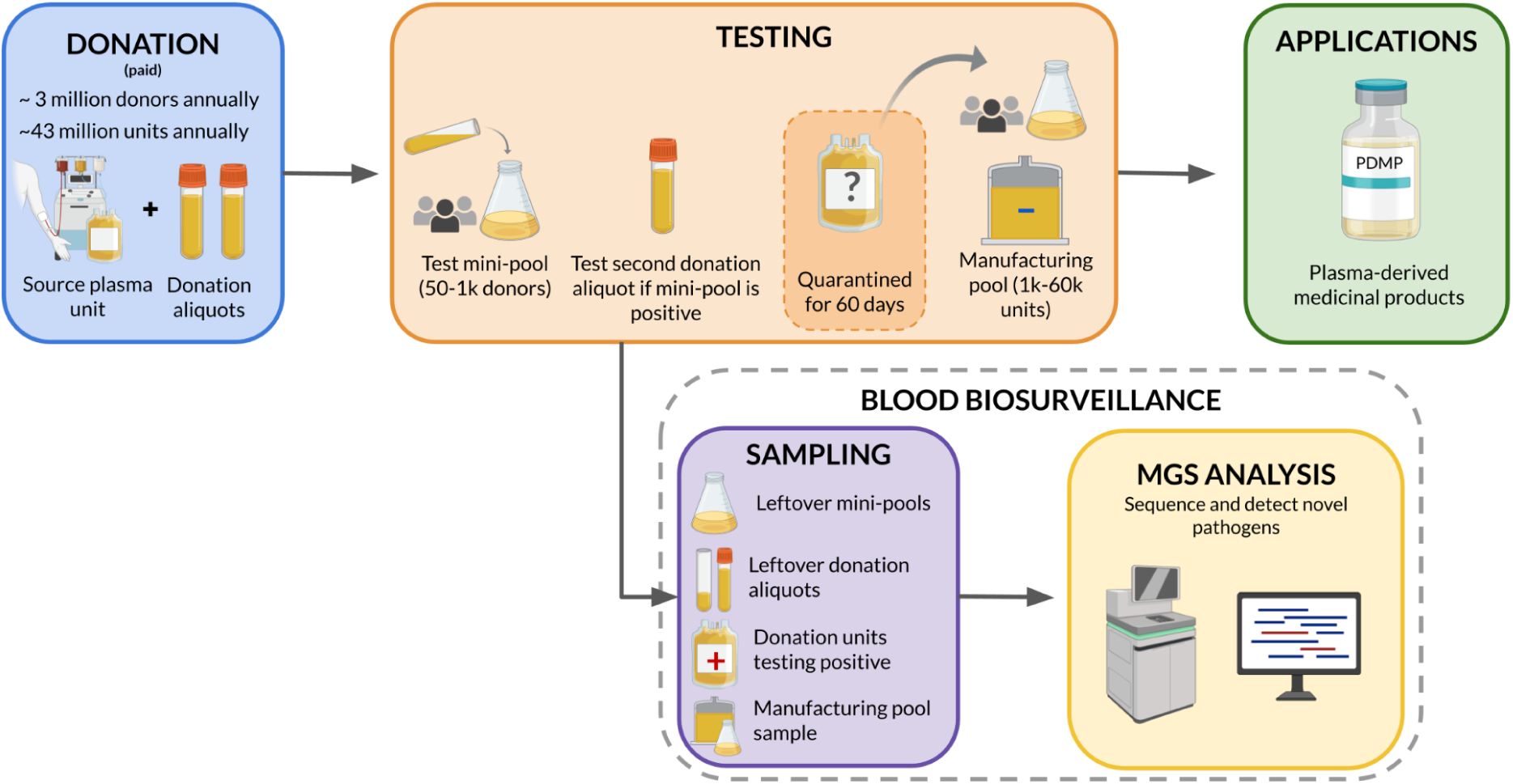
Sampling strategies in the plasma industry for metagenomic sequencing (MGS)-based biosurveillance. The standard plasma donation and testing process creates several sample streams, including leftover donation aliquots, mini-pools, transfusion-transmitted infections (TTI)-positive units, and manufacturing pools, that can be repurposed for MGS and pathogen-agnostic detection with minimal operational disruption.

**Figure 3.**
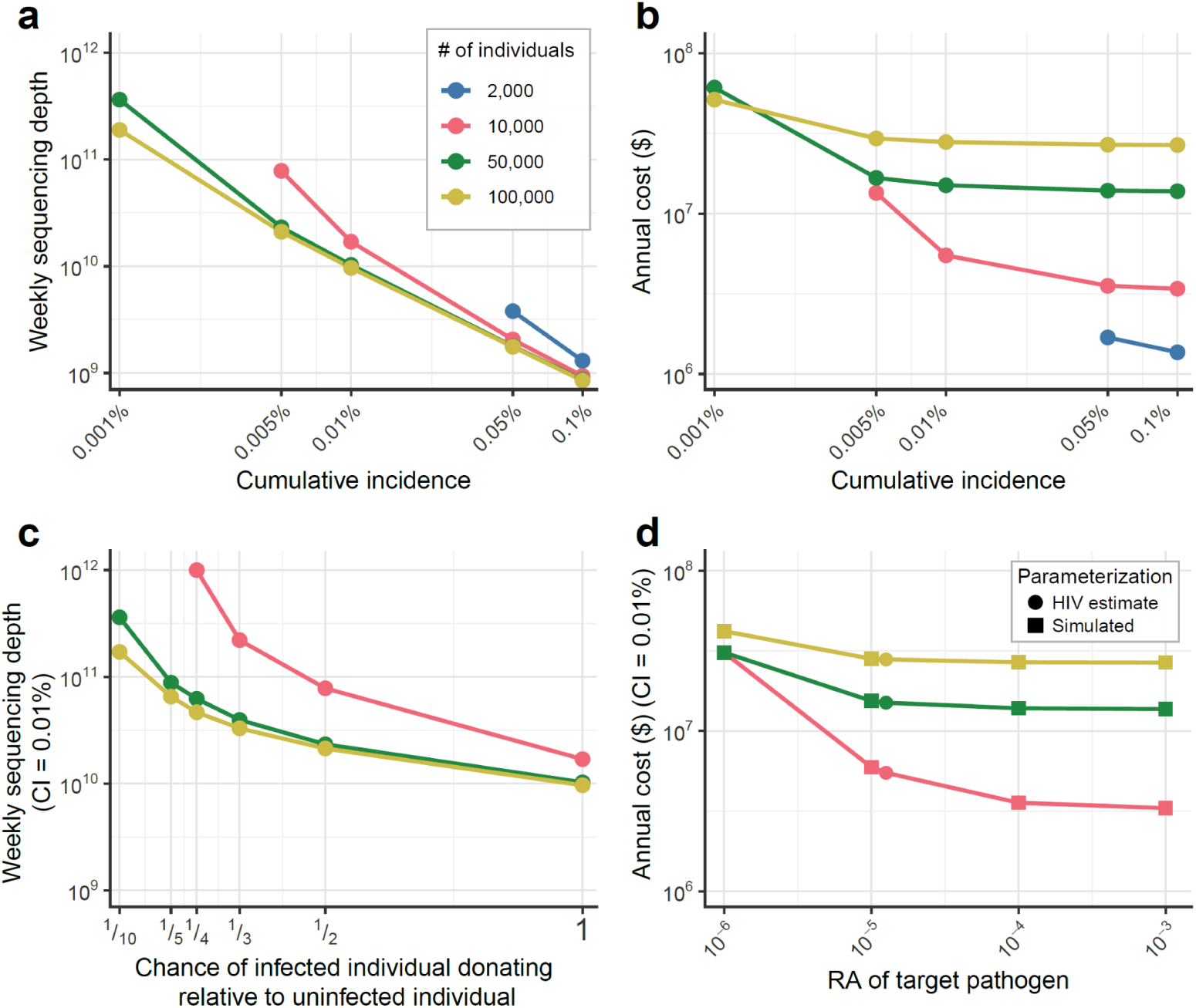
Modeling the cost of an Metagenomic Sequencing (MGS)-based blood-supply early pathogen detection system. (a) Required weekly sequencing depth and (b) corresponding estimated annual cost to achieve detection across different cumulative incidence targets, based on the number of individuals sampled per week. (c) Relationship between the chance of an infected individual donating relative to uninfected individuals, holding the detection target constant at 0.01% cumulative incidence. (d) Relationship between pathogen relative abundance and annual cost, holding the detection target constant at 0.01% cumulative incidence.

We identify a cumulative incidence of 0.01% (approximately 34,000 infections 2.3 years into the outbreak) as a practical baseline target. Achieving this level of detection would require sampling 10,000 individuals and sequencing around 17 billion reads per week, throughput achievable on a single Illumina NovaSeq X flow cell, at an estimated cost of $5.5 million annually (cost breakdown in Table 1). Estimated annual costs ranged from $3.5 million to $26.2 million as assumed associated mean relative abundance of pathogen reads in infected individuals was varied an order of magnitude above and below the central scenario in Table 1, respectively. Detection at a cumulative incidence of 0.01% would substantially improve upon symptom-based clinical surveillance, which, in the case of HIV, only occurred after nearly a decade of undetected spread and millions of infections.

**Table 1.** Estimated annual cost breakdown for detecting a novel HIV-like pathogen at 0.01% cumulative incidence. Model assumes a weekly sampling cohort of 10,000 individuals.

| **Cost component** | **Annual cost** | **Percentage** |
| --- | --- | --- |
| Sample acquisition | \$2,600,000 | 47.4% |
| Sequencing | \$2,205,000 | 40.2% |
| Labor | \$650,000 | 11.8% |
| Sample processing | \$26,000 | 0.4% |
| Data storage & processing | \$6,700 | 0.1% |
| Total | \$5,487,000 | |

The system’s cost-effectiveness could be further improved by optimizing sample preparation to increase the relative abundance of target nucleic acids, thereby reducing sequencing requirements. For example, a tenfold increase in the pathogen’s relative abundance (from µ≈10^-^⁵ to 10^-^⁴) would lower the annual cost for the 0.01% detection target from $5.5 million to $3.6 million, as shown in Figure 3d. However, such gains show diminishing returns, as the principle limitation eventually shifts from sequencing depth to the rarity of infected individuals within the weekly sampling cohort.

Several limitations should be noted. First, our model ignores sub-national heterogeneity in early epidemic spread — localized outbreaks could be either easier to detect (if sampling happens to be concentrated in affected areas) or harder (if spread occurs in areas with more limited sampling). Second, our results are sensitive to the assumed relative abundance of the pathogen in infected donors – a parameter that is likely to vary considerably across pathogens and stages of infection. Third, the model assumes that a novel pathogen can be reliably identified from metagenomic reads once a sufficient number have been sequenced; in practice, the bioinformatic challenge of detecting a truly unknown organism without a reference genome remains a significant open problem.

## 4. Viral targets in the blood supply

The effectiveness of MGS-based blood supply pathogen monitoring depends on which emerging viruses can be detected among predominantly asymptomatic donors. Viremia, defined here as the presence of viral nucleic acids or, in some contexts, infectious particles in the bloodstream, is a prerequisite for detection by MGS. Patterns of viremia vary substantially across viruses and human hosts, determining the suitability of blood-based monitoring for different pathogen types and informing MGS strategy.

Blood supply biosurveillance is well-suited for detecting viruses that cause asymptomatic viremia. These include pathogens like human parvovirus B19, which can produce substantial viremia before symptom onset,^55^ and viruses like HIV, HBV, HCV, and cytomegalovirus (CMV), which evade immune clearance to establish persistent or latent infections in blood cells or nearby tissues. For instance, HIV RNA remains detectable in plasma during clinical latency,^56^ and HBV produces circulating viral DNA in asymptomatic individuals with persistent liver infections.^57^ MGS studies of blood from healthy or asymptomatic individuals, including blood donors, reveal a “healthy blood virome” dominated by persistent DNA viruses such as anelloviruses and herpesviruses including EBV and CMV,^58–60^ consistent with their widespread seroprevalence. These findings suggest blood monitoring could provide an early warning for novel viruses with similar biological features, including strong immune evasion or a pre-symptomatic viremia, that might otherwise be missed by traditional syndromic surveillance.

Arboviruses, including dengue, Zika, and WNV, represent another promising target class. These infections often involve a high proportion of asymptomatic cases (up to 75-80%) that remain viremic and contribute to transmission.^61–63^ While plasma viremia may be brief, viral RNA for dengue, Zika, and potentially WNV can persist for weeks or even months in whole blood.^64,65^ This extended detectability makes blood-based surveillance well-suited for monitoring arboviruses, although it may lag detection by syndromic surveillance. Chikungunya virus also causes high viremia levels, though its asymptomatic viremic window appears shorter than that of other flaviviruses.

By contrast, blood-based MGS faces substantial limitations for detecting most respiratory and gastrointestinal viruses. Viremia in asymptomatic individuals with respiratory viruses such as influenza is exceedingly rare and typically reflects severe disease incompatible with blood donation.^66^ Large studies of blood donors have found influenza RNAemia to be virtually non-existent,^67–69^ with only two known instances of influenza RNA detection in asymptomatic individuals.^70,71^ SARS-CoV-2 is a partial exception: RNAemia was detected in 7.8-15% of presymptomatic blood donors who later developed COVID-19,^72^ although overall rates in pooled donor samples were estimated to be very low (< 0.01%).^27^ Gastrointestinal viruses, including rotaviruses and norovirus, remain largely restricted to the enteric tract, with no viremia detected even during active infection.^73^ Because of these biological constraints, blood monitoring will likely miss early outbreaks of respiratory or gastrointestinal viruses, and should therefore complement, rather than replace, other systems like wastewater and clinical surveillance, which may in turn be less suitable for monitoring blood-borne pathogens.

Effective MGS detection also depends on sample preparation strategies tailored to the biology of viral infection in blood. Viral nucleic acids are vastly outnumbered by host material; human genomic DNA can constitute the vast majority of sequence reads from whole blood, reducing both MGS cost-efficiency and sensitivity. Moreover, viral nucleic acids are present in different blood compartments – within blood cells (e.g., episomes, integrated proviruses, or replicating viruses), as cell-free virions in plasma, or as cell-free nucleic acids (cfNA) in plasma. The optimal sample type for detection thus varies by pathogen. While no amount of optimization can detect a virus that is not present, various preparation strategies can enrich the viral signal in viremic samples. These include filtration to remove host cells,^59^ nuclease treatment to enrich for encapsulated virions, and targeted depletion of human sequences to increase the fraction of viral reads.^74^ Balancing these tradeoffs is an important area for further research to maximize the cost-effectiveness and sensitivity of MGS-based blood surveillance.

## 5. Privacy and regulatory considerations

Large-scale public health monitoring of the U.S. blood supply during the COVID-19 pandemic established a strong regulatory and privacy framework for implementing MGS-based pathogen surveillance. During the pandemic, a CDC-led program analyzed over 1.4 million blood donor specimens, with studies approved as either “non-research public health surveillance”^28^ or as research not involving human subjects.^29,75^ These determinations under the Common Rule were based on the use of anonymized data and standard donor consent, which allows residual samples to be used for research. This precedent suggests that a properly designed MGS monitoring system using de-identified residual samples could proceed without requiring additional donor consent or full IRB review. When MGS findings warrant confirmatory testing on individual aliquots within a flagged pool, blood collection and testing organizations can use the same well-established and secure traceback infrastructure that already supports TTI testing.

While MGS warrants special consideration because it sequences human nucleic acids alongside pathogen sequences, several safeguards mitigate inadvertent re-identification risk. Blood collection organizations already perform de-identification in accordance with the Health Insurance Portability and Accountability Act (HIPAA), removing all Protected Health Information (PHI) before samples are released for analysis. Sequencing pooled materials, such as residual mini-pools from TTI testing, further protects privacy by masking individual contributions. Data management best practices add an additional layer of protection through controlled-access systems and bioinformatic pipelines that automatically remove human sequences before downstream analysis. Emerging experimental technologies can further enhance privacy by physically depleting human nucleic acids from a sample prior to sequencing.^74^

Together, these genomic and procedural safeguards provide robust privacy protection while maintaining regulatory compliance under frameworks such as the Genetic Information Nondiscrimination Act (GINA), which protects heritable genetic information but excludes microbial analysis. As a result, MGS-based monitoring of de-identified blood supply samples is well supported by existing ethical and regulatory precedent.

## 6. Conclusion

MGS-based monitoring of the U.S. blood supply offers a tractable and high-leverage addition to national early warning capabilities. Residual minipools from existing TTI testing can be sequenced and analyzed within days of collection, enabling rapid detection of emerging pathogens that produce viremia in asymptomatic donors. The approach is particularly suited to novel blood- and vector-borne viruses with long asymptomatic periods and immune-evasive properties, characteristics that delayed recognition of HIV, HCV, and other outbreaks during their early spread. Our cost analysis suggests that an effective nationwide implementation could be achieved with an annual investment of approximately $5.5 million, substantially improving on symptom-based surveillance and potentially identifying stealthily spreading threats months or years before they would otherwise be recognized.

Such a system would complement rather than replace existing surveillance infrastructure. Targeted TTI testing protects blood safety through sensitive assays for known pathogens, and other programs such as the CDC’s Traveler-based Genomic Surveillance and National Wastewater Surveillance System monitor different populations and matrices. Blood-supply MGS extends this layered network to a sample type that is uniquely positioned to detect novel blood-and vector-borne pathogens, while drawing on infrastructure that already exists. Three decades of NHLBI- and FDA-funded blood donor research programs, including expanding serosurveillance work in partnership with the CDC, have built the multi-center networks, sample processing workflows, and data sharing systems that the proposed system would draw on.

Implementation can begin with the operational architecture developed here and extend over time as MGS matures. Near-term deployment is feasible with current technology, current regulatory frameworks, and existing partnerships between blood collection organizations, public health agencies, and research institutions. As the technology matures and validation evidence accumulates, MGS could be integrated more deeply into TTI testing workflows themselves, further strengthening transfusion safety alongside the early-warning capabilities. Although this paper focuses on the U.S., the same framework is broadly applicable to countries with comparable blood collection infrastructure. National investment in this capability would be modest relative to the costs of unchecked spread of a novel pathogen due to delayed detection, and the infrastructure is in place to begin now.

## Data Availability

All code and data to reproduce the cost modeling analysis are publicly available at https://github.com/naobservatory/blood-cost-modeling.

https://github.com/securebio/blood-cost-modeling

## 7. Acknowledgements

We thank Dan Rice for assistance with the modeling, Natalia Wewior for sharing her expertise on regulatory aspects of metagenomic sequencing, and Eric Delwart, Will Bradshaw, Simon Grimm, and Nils Justen for their feedback on the manuscript. Figure 1 and 2 used graphic resources from BioRender: Justen, L. (2026) https://BioRender.com/62jcam3.

## 8. Author contributions

**Conceptualization:** L.J.J., M.C.L., M.P.B., K.M.E., P.C.S. **Methodology:** L.J.J., H.B., C.W. **Software:** H.B., C.W. **Validation:** L.J.J. **Formal analysis:** L.J.J., H.B., C.W. **Investigation:** L.J.J., H.B. **Writing – original draft:** L.J.J., H.B. **Writing – review & editing:** C.W., D.C.-B., M.C.L., M.P.B., K.M.E., P.C.S. **Visualization:** L.J.J., H.B. **Supervision:** D.C.-B., K.M.E., P.C.S. **Project administration:** L.J.J. **Funding acquisition:** K.M.E., P.C.S.

## 11. Code availability

All code and data to reproduce the cost modeling analysis are publicly available at https://github.com/securebio/blood-cost-modeling.

