## appendix for "Leveraging the U.S. blood supply to detect emerging viral threats"

### Appendix — Modeling the cost of a metagenomic sequencing-based blood supply early pathogen detection system

April 2026

**3 Estimating cost** **5**

#### 1 Introduction

We developed a cost modeling framework to estimate the annual expense of operating a metagenomic sequencing (MGS)-based, blood supply pathogen early detection system. Our framework stipulates that an effective blood monitoring system should be able to detect an outbreak of a pathogen before a certain fraction of the population has been infected – a cumulative incidence target. For a given cumulative incidence target, as well as a set of pathogen properties, and system design choices, we model the sequencing depth (number of reads) needed to detect the pathogen; depending on the configuration, detection may not be possible at a particular cumulative incidence. To estimate the annual cost of the system, we translate the sequencing depth requirements for detection into sequencing costs, and combine this with estimates for sampling costs, compute, and labor. Taken together, the model represents a modular and flexible system for modeling MGS-based detection through the blood supply.

Modeling the necessary sequencing depth for detection is the most complex and important aspect of our approach. It combines four sub-modules: an outbreak module, shedding dynamics module, sampling and sequencing module, and detection module, drawing on insights from past work [1, 2]. Collectively, these modules model how the pathogen spreads through the population and infects blood donors, how blood donors are sampled from in our modeled system, how infected individuals shed the pathogen into blood, and how MGS reads out sequences from the target pathogen and triggers detection. The way that each of these processes are formalized and parameterized requires simplifications and assumptions, which we try to list and make explicit.

Using the model, we evaluate the cost of detecting a hypothetical novel HIV-like pathogen spreading through the population with MGS and a blood sampling system similar in design to past CDC-led efforts during COVID-19 to monitor seroprevalence [3–5]. We choose HIV as an example of a pathogen which can spread for years before distinguishable symptoms, has historically spread through the blood supply undetected, and would evade many of our current symptom-based early detection systems. Below, we describe the model as well as our HIV-like pathogen parameterization.

Code to reproduce the model and figures is available here: `https://github.com/securebio/blood-cost-modeling`

#### 2 Modeling sequencing requirements

##### 2.1 Outbreak

The outbreak module simulates the spread of a pathogen through a population. It determines how many people are infected each week and cumulatively over time, and when the epidemic reaches specific cumulative incidence thresholds that serve as our detection targets. When we go to sample blood donors for sequencing in the sampling and sequencing module, the outbreak module determines the fraction of contributing individuals who are infected.

We model the epidemic using a deterministic renewal equation, a standard framework for early outbreak dynamics [6, 7]. New infections in week $t$ are given by:

$$I(t) = R_0 \sum_{k=1}^{\infty} I(t-k)\, w(k) \tag{1}$$

where $R_0$ is the basic reproduction number and $w(k)$ is the generation time distribution, representing the probability that a secondary infection occurs $k$ weeks after the primary infection. We assume uniform infectiousness over a finite infectious period of $D$ weeks, giving $w(k) = 1/D$ for $k = 1, \dots, D$ and $w(k) = 0$ otherwise. Under this assumption, the renewal equation simplifies to:

$$I(t) = \frac{R_0}{D} \sum_{k=\max(1,\,t-D)}^{t-1} I(k) \tag{2}$$

The relationship between $R_0$, $D$, and the asymptotic exponential growth rate $r$ is determined by the Euler–Lotka equation:

$$R_0 = \frac{D}{\sum_{k=1}^{D} e^{-rk}} \tag{3}$$

which we use to derive $R_0$ from a specified growth rate and infectious period.

To parameterize the outbreak module, we set the asymptotic growth rate to $r = 0{\cdot}0155$ per week, corresponding to a doubling time of approximately 45 weeks (0·86 years), based on Worobey and colleagues' analysis of early

HIV-1 subtype B spread among men who have sex with men in the United States during the 1970s [8]. For the baseline analysis, we set $D = 12$ weeks, equal to the shedding duration, yielding $R_0 \approx 1 \cdot 10$ via the Euler–Lotka equation. We additionally explore $D = 52$ weeks as a sensitivity analysis, representing a pathogen with a longer infectious period and a correspondingly higher $R_0 \approx 1 \cdot 47$ at the same growth rate. Because $r$ is held constant and the shedding duration is defined independently of $D$, the number of individuals actively shedding at any given cumulative incidence threshold is approximately the same regardless of $D$, and so $D$ has limited influence on detection requirements for cumulative-incidence-based targets. We note, however, that $D$ would influence other later-stage epidemic dynamics (such as final size) through its effect on $R_0$, though these are of less relevance here given our focus on early detection when spread has been minimal. We set $I_1 = 100$ initial infections to ensure the outbreak persists beyond stochastic fluctuations that could terminate smaller seed populations [9, 10].

We evaluate detection across cumulative incidence thresholds ranging from 0·0001% to 0·1% of the U.S. population ($N = 340$ million), equivalent to 340 to 340,000 infections before detection. This range spans a range of potential system objectives: detection at 0·0001% (340 infections) could enable geographic containment, while even 0·1% (340,000 infections) could yield substantially earlier detection of an HIV-like pathogen than symptom-based surveillance, potentially accelerating medical countermeasure development by months.

##### 2.2 Shedding dynamics

The shedding dynamics module determines how much pathogen genetic material is present in the blood of infected individuals over time. It assumes that infected individuals shed the pathogen into blood for a particular amount of time post-infection, $\epsilon$, and at particular rate i.e., the relative abundance of the pathogen in MGS data from an infected individual, $\mu$. For simplicity, we assume that the shedding rate remains constant over the shedding period, and that shedding period does not vary across individuals, though this obscures more complex host-pathogen-specific dynamics. We note that the shedding duration $\epsilon$ is conceptually distinct from the infectious period $D$ used in the outbreak module: $\epsilon$ governs how long an individual contributes detectable pathogen reads to blood samples, while $D$ governs how long an individual contributes to onward transmission. In our baseline parameterization these are set equal ($\epsilon = D = 12$ weeks), but the model allows them to vary independently. We define the number of individuals actively shedding in a given week, $t$, denoted $S(t)$, as the sum of all infected individuals during the previous $d$ weeks:

$$S(t) = \sum_{k=\max(1,t-\epsilon)}^{t} I(k).$$

For HIV viremia, we assume each infected individual sheds the pathogen continuously at a constant rate over a 12-week period following infection. Although actual HIV shedding varies considerably over time, our approach focuses on the acute infection phase, characterized by high and relatively consistent shedding, after which viral loads typically decline before eventually increasing again with progression to AIDS [11].

We parameterize the pathogen shedding rate $\mu$ drawing from using data from Piantadosi and colleagues [12]. Specifically, we calculate the arithmetic mean relative abundance of HIV (HIV reads divided by total reads) in plasma from three individuals within two weeks of infection, $\mu = 1 \cdot 26 \times 10^{-5}$. To account for uncertainty arising from this limited sample size, as well as variation due to sample processing protocols, sample type (whole blood versus plasma), and individual- and pathogen-specific shedding dynamics [13], we explore values of $\mu$ ranging from $10^{-8}$ to $10^{-3}$ in the results section.

##### 2.3 Sampling and sequencing

The sampling and sequencing module determines the sampling of infected and non-infected donors from the population and the resulting number of sequences read from the target pathogen for a given sequencing effort.

We model the number of infected, actively shedding individuals captured in a weekly sample as a binomial distribution: $A(t) \sim \text{Binomial}\left(P, \frac{S(t)}{N}\right)$, where $A(t)$ is the number of sampled shedders, $S(t)$ represents the number of individuals actively shedding the pathogen in week $t$, and $P$ represents the total number of individuals sampled. In practice, infected individuals may be less likely to donate blood than uninfected individuals, for example due to feeling unwell. To account for this, we introduce a relative risk parameter $\rho$ representing the chance of an infected individual donating relative to an uninfected individual. The effective number of shedding donors available for sampling in week $t$ becomes $S(t) \times \rho$, and the binomial sampling probability is adjusted accordingly: $A(t) \sim \text{Binomial}\left(P, \frac{S(t)\times\rho}{N}\right)$. In our baseline scenario we set $\rho = 1$, assuming infection status does not influence donation probability, and explore values ranging from $\rho = 0 \cdot 1$ to $\rho = 1$ as a sensitivity analysis.

From this we calculate the expected pathogen reads yielded by a given sequencing effort (number of sequences, $D$) at a given time in the outbreak as:

$$E[R(t)] = \frac{A(t)}{P} \times \mu \times D$$

This formulation assumes all $P$ individuals' samples are combined into a single batch for sequencing. In practice, samples might be obtained as pre-existing mini-pools (e.g., 16-person pools from routine blood screening) or individual aliquots that could be combined into custom pool sizes. While our model treats these scenarios equivalently in terms of $P$ contributing individuals, the actual implementation would need to consider that different pooling strategies could affect both costs and detection sensitivity.

The term $\frac{A(t)}{P}$ represents the fraction of infected, shedding individuals in our pooled sample, which appropriately accounts for dilution when combining infected and uninfected samples. For example, if 10 out of 1,000 sampled individuals are infected, the effective relative abundance in the pool becomes $\frac{10}{1000} \times \mu = 0 \cdot 01$, reflecting a 100-fold dilution compared to an individual infected sample.

To capture the biological variability inherent in viral shedding, we model actual pathogen reads using a negative binomial distribution – a choice well-supported for overdispersed sequence count data [14, 15]. We parameterize the mean of the distribution based on $E[R(t)]$ and model dispersion based on $\frac{A(t)}{CV^2}$, with $CV^2 = 0 \cdot 69$ estimated from the coefficient of variation squared of relative abundances from same HIV-data used to estimate $\mu$ [12]. This partially accounts for the variation in viral loads between individuals and over time.

In parameterizing our sampling system, we use estimates from the whole blood transfusion system rather than source plasma, because the former has demonstrated utilization for population-level pathogen monitoring during COVID-19 RESPONSE [3].

We model weekly sampling as independent selection with replacement from the total population of 340 million. This simplification assumes we sample from the entire U.S. population rather than the approximately 6·6 million annual blood donors [16], who generally skew healthier, older, whiter, and wealthier than average while excluding high-risk groups through eligibility screening [17, 18]. While this assumption is reasonable for a pathogen that spreads uniformly through the population, it will yield overly optimistic cost estimates for outbreaks concentrated in populations underrepresented in the donor pool (e.g., children), whether due to exclusion or other factors.

Each week, we randomly select $P$ contributing individuals, where $P$ ranges from 2,000 to 100,000. We consider $P = 10{,}000$ as a realistic baseline given national blood monitoring efforts during COVID-19. For instance, Bakkour and colleagues [5] performed minipool testing for SARS-CoV-2 RNA early in the pandemic, processing on average around 620 minipools each week, representing approximately 9,000 weekly individual donations. A U.S. national COVID-19 serosurveillance program in partnership with CDC processed around 38,000 blood donations per week [3]. On the upper bound, the whole blood transfusion system processed 11·5 million annual donations in 2021 [16], suggesting a baseline average of around 200,000 weekly donations.

Our sampling and sequencing model makes several simplifying assumptions. We aggregate all U.S. samples into a single weekly sequencing batch, ignoring geographic stratification that could improve early detection in localized outbreaks. We assume one sequencing run per week, though operational systems might sequence more or less frequently. We ignore delays between when the samples are collected from individuals, when they are available for processing and sequencing, and when the data is analyzed for pathogen detection, assuming that this can be reduced to less than a week. We also treat mini-pools and individual samples equivalently in terms of pathogen detection probability, though pooling dilution effects could reduce sensitivity.

##### 2.4 Detection

The detection module determines the minimum sequencing depth required to detect the target pathogen at a certain target cumulative incidence. We stipulate that detection occurs when the number of sequencing reads generated by our proposed monitoring system attributable to the target pathogen surpasses a certain read threshold, $\theta$.

We use a Monte Carlo simulation approach [19, 20] to model detection to account for randomness involved in pathogen spread, individual shedding rate, and sampling of infected donors. In our simulation framework, we specify a given weekly sequencing depth $D$ and a target cumulative incidence, and then simulate an exponentially growing outbreak week by week with stochastic donor sampling until cumulative pathogen reads surpass the detection threshold.

$$\sum_{t=1}^{T_{detect}} R(t) \geq \theta$$

To identify the optimal sequencing depth for a given cumulative incidence target, we implement a binary search algorithm constrained between $10^4$ reads (minimal practical depth) and $10^{12}$ (one trillion read pairs, out of range for current monitoring systems). For each candidate depth, we conduct 500 Monte Carlo simulations to estimate $P(\text{detect before } X\% \text{ incidence})$. If this probability falls below 95%, we increase sequencing depth; if it exceeds 95%, we decrease it. The binary search continues until the upper and lower bounds differ by less than 10,000 reads, yielding the minimal sequencing depth:

$D^* = \min\{D : P(\text{detect before } X\% \text{ incidence}) \geq 95\%\}$

Detection timing varies considerably across simulations due to the stochastic nature of both epidemic spread and sampling. In some cases—particularly when targeting extremely low incidence (e.g., 0·0001%) with small sample sizes or insufficient sequencing depth—simulations never accumulate enough pathogen reads to trigger detection before the target incidence is reached. We call these "non-convergent" solutions, which are especially likely with our HIV-like pathogen's slow spread (doubling time approximately 45 weeks) that limits infected individuals available for early sampling.

In parameterizing the model for detection of a novel HIV-like pathogen, we set $\theta = 100$ reads. In practice, the number of reads required for detection depends on the bioinformatic approach used (e.g., homology to known pathogens, anomaly detection relative to a well-characterized background, or monitoring for exponential growth) and its corresponding sensitivity. A full genome assembly is not a prerequisite for an initial detection; a modest number of reads can be sufficient to flag a sample as suspicious and trigger follow-up characterization using more sensitive approaches.

#### 3 Estimating cost

To estimate the cost of a MGS-based blood supply early pathogen detection system, we translate the sequencing depth requirements from the modeling framework into a cost of sequencing, and combine it with other operating expenses such as sample acquisition, labor, and data processing and storage. Taken together, we arrive at a model the returns the expected annual cost for detecting an outbreak using MGS of the blood supply at different cumulative incidence targets.

We model annual cost as:

$$C_{\text{annual}} = \sum_{w=1}^{52} \left(C_{\text{weekly\_ops}} + \left(w \times C_{\text{storage\_unit}}\right)\right) + C_{\text{labor}}$$

where $C_{\text{weekly\_ops}} = (C_{\text{samp}} \times P) + C_{\text{samp\_proc}} + D \times (C_{\text{seq}} + C_{\text{data\_proc}})$ represents the constant weekly operational costs for sample acquisition, processing, sequencing, and data processing at a fixed number of sampled individuals, $P$, and sequencing depth in billions of reads, $D$; and $C_{\text{storage\_unit}} = D \times C_{\text{data\_store}}$ represents the cost to store a single week's batch of new data for one week. The term $w \times C_{\text{storage\_unit}}$ calculates the storage cost for a given week, accounting for the data from week 1, week 2, ..., all the way up to the current week, $w$.

Sample collection costs $C_{\text{samp}}$ represent the per-individual cost of accessing blood samples for pathogen monitoring. Our research into large-scale, real-world biosurveillance programs suggests a plausible range of \$1·00 to \$20·00 per individual, with a central estimate of \$5·00. This range is grounded in data from the CDC's COVID-19 surveillance efforts. The lower bound is informed by the CDC's \$46·4 million prime contract with Vitalant (75D30120C08170) [21], which yielded an all-inclusive cost of approximately \$6·44 per sample for the entire serosurveillance service; the marginal cost for sample acquisition alone would be a fraction of this total. The upper bound is supported by a CDC subcontract (11781ARC001) that paid the American Red Cross approximately \$19·34 per individual for a more comprehensive follow-up effort that included sample acquisition, processing, repository work, and donor surveys.

Batch processing costs $C_{\text{samp\_proc}}$, which encompass nucleic acid extraction and library preparation, are assumed to be a fixed cost of \$500 per week using data from MIT BioMicroCenter pricing [22], though actual costs vary by platform and protocol.

For sequencing costs, we assume $C_{\text{seq}} = \$2{,}500$ per billion read pairs based on Illumina NovaSeq X pricing, following estimates from Grimm and colleagues [1]. A fixed per-read cost represents simplification of the discontinuous pricing structures involved with sequencing; cost per read can vary substantially depending on sequencing volumes, with pricing often being at the level of a flowcell or even lanes within a flowcell.

For MGS data processing $C_{\text{data\_proc}}$, we estimate a cost of \$1 per billion read pairs processed. For data storage $C_{\text{data\_store}}$, we estimate that a billion read pairs costs \$0·25 per week to store. These estimates are derived from real-world MGS pathogen monitoring pilots carried out by the SecureBio Detection team [23].

Finally, for labor, we consider an annual cost of \$650,000, consisting of salaries for a lab manager, lab technician, and two bioinformaticians, based on NAO labor cost estimates.

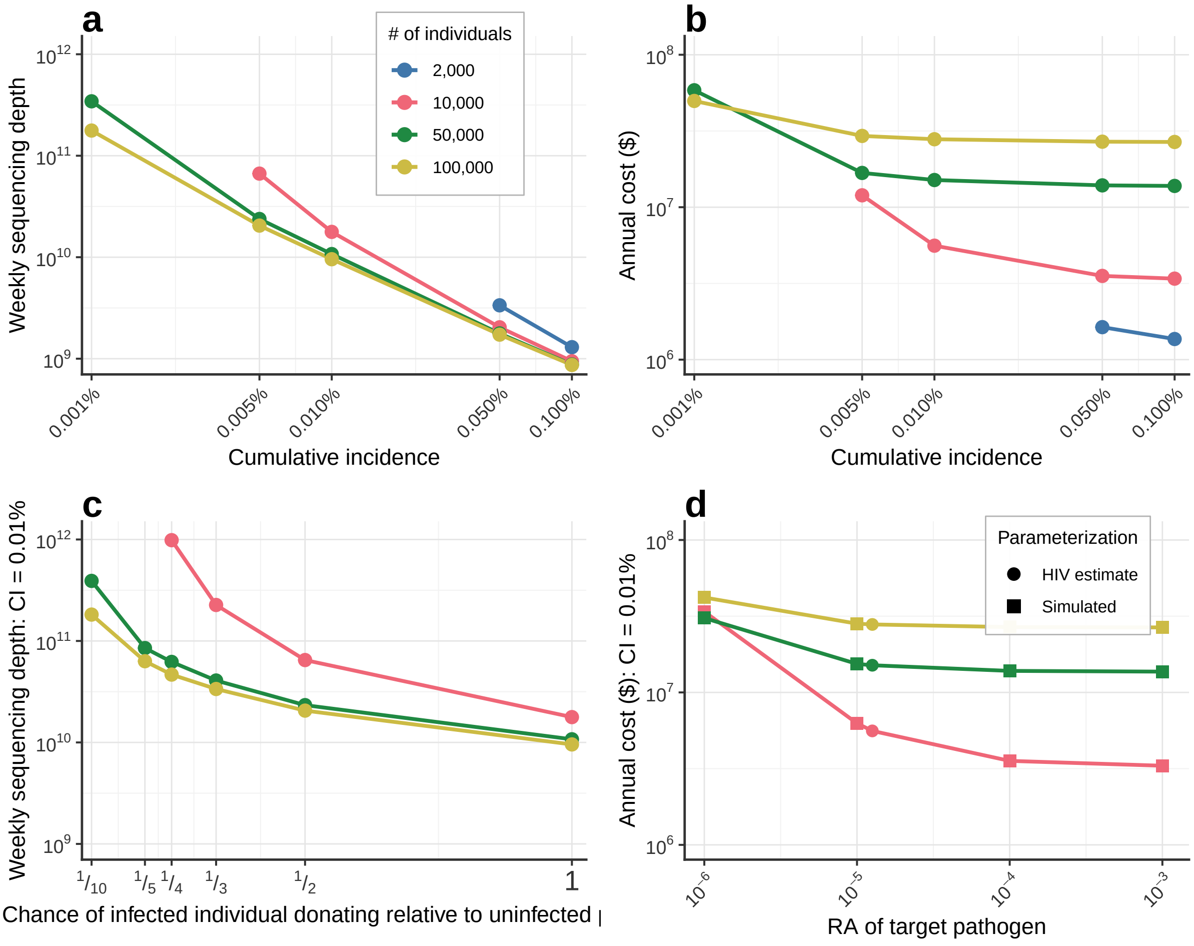

**Supplementary Figure 1: Sensitivity analysis showing the effect of infectious period duration on detection requirements.** Results are shown for $D = 52$ weeks, compared to the baseline $D = 12$ weeks in Figure 3 of the main text. Because the asymptotic growth rate $r$ is held constant and shedding duration is defined independently of $D$, detection requirements are largely unchanged across values of $D$.
